# Risk factors for adverse clinical outcomes in patients with COVID-19: A systematic review and meta-analysis

**DOI:** 10.1101/2020.05.13.20100495

**Authors:** Vanesa Bellou, Ioanna Tzoulaki, Evangelos Evangelou, Lazaros Belbasis

**Affiliations:** Department of Hygiene and Epidemiology, University of Ioannina Medical School, Ioannina, Greece; Department of Epidemiology and Biostatistics, School of Public Health, Imperial College London, London, United Kingdom

## Abstract

**Importance:** COVID-19 is a clinically heterogeneous disease of varying severity and prognosis. Clinical characteristics that impact disease course could offer guidance for clinical decision making and future research endeavors and unveil disease pathways.

**Objective:** To examine risk factors associated with adverse clinical outcomes in patients with COVID-19.

**Data sources:** We performed a systematic review in PubMed from January 1 until April 19, 2020.

**Study selection:** Observational studies that examined the association of any clinical characteristic with an adverse clinical outcome were considered eligible. We scrutinized studies for potential overlap.

**Data extraction and synthesis:** Information on the effect of clinical factors on clinical endpoints of patients with COVID-19 was independently extracted by two researchers. When an effect size was not reported, crude odds ratios were calculated based on the available information from the eligible articles. Study-specific effect sizes from non-overlapping studies were synthesized applying the random-effects model.

**Main outcome and measure:** The examined outcomes were severity and progression of disease, admission to ICU, need for mechanical ventilation, mortality, or a composite outcome.

**Results:** We identified 88 eligible articles, and we performed a total of 256 meta-analyses on the association of 98 unique risk factors with five clinical outcomes. Seven meta-analyses presented the strongest epidemiological evidence in terms of statistical significance (P-value <0.005), between-study heterogeneity (I^2^ <50%), sample size (more than 1000 COVID-19 patients), 95% prediction interval excluded the null value, and absence of small-study effects. Elevated C-reactive protein (OR, 6.46; 95% CI, 4.85 – 8.60), decreased lymphocyte count (OR, 4.16; 95% CI, 3.17 – 5.45), cerebrovascular disease (OR, 2.84; 95% CI, 1.55 – 5.20), chronic obstructive pulmonary disease (OR, 4.44; 95% CI, 2.46 – 8.02), diabetes mellitus (OR, 2.04; 95% CI, 1.54 – 2.70), hemoptysis (OR, 7.03; 95% CI, 4.57 – 10.81), and male sex (OR, 1.51; 95% CI, 1.30 – 1.75) were associated with risk of severe COVID-19.

**Conclusions and relevance:** Our results highlight factors that could be useful for prognostic model building, help guide patients’ selection for randomized clinical trials, as well as provide alternative treatment targets by shedding light to disease pathophysiology.

## 1 INTRODUCTION

In December 2019, a cluster of pneumonia cases was reported in Wuhan, China and subsequent epidemiological tracking identified a novel coronavirus (Severe Acute Respiratory Syndrome-Coronavirus-2, SARS-CoV-2) as the cause.^1^ Two other coronaviruses, causing Severe Acute Respiratory Syndrome and Middle East Respiratory Syndrome, were responsible for epidemics originated in Asia in the past. The SARS-CoV-2 has spread across all continents since December and caused a public health crisis.^2^ As of late April 2020, there has been more than 3 million cases and more than 200,000 deaths according to WHO.^3^ A significant percentage of patients have severe or critical disease and require intensive care management.^4^ Currently there is only one approved medication for Coronavirus Disease 2019 (COVID-19), and many questions on clinical management remain unanswered.^5^ To this end, more than 300 clinical trials are being conducted to find an effective treatment.^2^

The ongoing public health emergency necessitates the discovery of reliable prognostic factors to help guide clinical decision making tailored to the patient characteristics. The identification of patient characteristics that are associated with adverse clinical events could provide alternative therapeutic targets as well as improve the design and analysis of future clinical trials.^6,7^ Also, the deciphering of risk factors having a detrimental effect on disease course could offer novel insights on molecular pathways of the disease.

There is a growing body of published observational studies on COVID-19 patients, examining clinical characteristics and diagnostic features of the disease. However, there is no published effort that systematically summarizes the effect of all the examined clinical variables on the observed adverse clinical outcomes in patients diagnosed with COVID-19. Our study aims to fill this gap by systematically mapping all the available evidence on the association of various clinical variables with risk for multiple clinical outcomes in patients with COVID-19. We also examine for potential biases and we extensively search for potential overlapping studies that distorts the existing evidence.

## 2 METHODS

### 2.1 Search strategy and eligibility criteria

We followed the MOOSE (Meta-analyses of Observational Studies in Epidemiology) guideline for conducting and reporting our meta-analysis.^8^

We systematically searched PubMed from January 1, 2020 to April 19, 2020 to identify observational studies examining risk factors of adverse clinical outcomes in patients diagnosed with COVID-19. We used the following search strategy: “coronavirus”[All Fields] OR “SARS-CoV-2”[All Fields] OR “cov”[All Fields] OR “2019-SARS-CoV-2”[All Fields] OR “COVID-19”[All Fields] OR “SARS-CoV-2”[All Fields]. This is the search algorithm used by LitCovid, which is a curated literature hub for tracking up-to-date scientific information about SARS-CoV-2.^9^ The literature search was performed by two independent researchers (VB, LB). We considered demographic and anthropometric individual characteristics, biomarkers measured in serum or other biological fluids, symptoms, clinical signs, medical history and comorbid diseases, patterns and findings in computed tomography of the chest. We focused on any clinical outcome in patients with COVID-19, such as mortality or worsening of clinical status.

We excluded observational studies that examined the differences in patients with pneumonia associated with COVID-19 and patients with community-acquired pneumonia. Furthermore, we excluded observational studies that examined risk factors for adverse clinical events in infections by other coronaviruses (e.g., Severe Acute Respiratory Syndrome, and Middle East Respiratory Syndrome). We excluded case-reports and case-series including less than 10 COVID-19 patients. We excluded observational studies published in languages other than English. Also, we scrutinized the geographic region, the involved hospitals, the time period of recruitment and the inclusion and exclusion criteria from each study to examine the presence of potentially overlapping populations.

### 2.2 Data extraction

Data extraction was performed independently by two researchers (VB, LB). From each eligible article, we extracted information on the first author, the year and the journal of publication, the geographic region and the involved hospital, the recruitment period, the examined risk factors, and the examined outcomes. For each risk factor, we extracted the reported measure of association (i.e., odds ratio, risk ratio, or hazard ratio) and the reported level of comparison. Whenever the level of comparison was not clearly presented for a measure of association, we did not extract it. Also, when such a measure was not reported, we extracted the number of exposed individuals in both cases and controls for binary exposures. We extracted information on risk factors measured at admission or initial clinical assessment to ensure temporality between the exposure and the observed outcome. For studies conducted in the same geographic region, we compared the hospitals, the recruitment periods, and the research teams to ensure that effect estimates from fully independent populations will be included in each meta-analysis. When multiple studies with samples from the same hospital examined the same association, we kept the study that had the largest sample size for the corresponding meta-analysis.

### 2.3 Statistical analysis

Whenever available, and in the absence of a reported effect size, we calculated the crude odds ratio (OR) and its standard error based on the reported 2 × 2 contingency table. When zero counts occurred in a cell of a 2 × 2 contingency table, we applied the Haldane-Anscombe correction.^10^ This is a method to avoid error in the calculation of odds ratio by adding 0.5 to all the cells of a 2 × 2 contingency table if any of the cell expectations would cause a division by zero. For associations examining risk factors for composite clinical outcomes, we performed a meta-analysis whenever the definition of the composite outcome was adequately similar across the different studies.

For each association, we estimated the summary effect estimate and its 95% confidence interval applying the random-effects model, because methodological heterogeneity was expected between the eligible studies.^11^ We used the DerSimonian and Laird estimator to estimate the between-study variance.^12^ A statistically significant effect was claimed at P-value <0.05. Also, between-study heterogeneity was quantified by the I^2^ metric.^13,14^ I^2^ ranges between 0% and 100% and is the ratio of between-study variance over the sum of the within-study and between-study variances. An I^2^ estimate larger than 50% was considered evidence for large heterogeneity, whereas an I^2^ larger than 75% was judged as very large heterogeneity.

For associations that were examined in at least 3 observational studies, we estimated the 95% prediction interval and we assessed the presence of small-study effects. 95% prediction interval further accounts the uncertainty for the effect that would be expected in a new study addressing that same association.^11^ We assessed whether there was evidence for small-study effects (i.e., whether smaller studies tend to give substantially larger estimates of effect size compared with larger studies) with the regression asymmetry test proposed by Egger and colleagues.^15^ The presence of small-study effects was based on a statistically significant Egger’s test at P-value <0.10 combined by a more conservative effect in the largest study of the meta-analysis compared to the effect in the random-effects meta-analysis.

We also examined which associations presented a highly significant effect at P-value <0.005, absence of large or very large between-study heterogeneity (i.e., I^2^<50%), 95% prediction interval excluding the null value, and sample size more than 1000 COVID-19 patients. The rationale for the use of a strict P-value threshold is based on current recommendations to avoid false-positive findings.^16–18^

Statistical analysis was performed on R, version 3.6.3.

## 3 RESULTS

We screened a total of 5924 articles, and we identified 88 eligible articles that fulfilled our eligibility criteria and were published between January 1, 2020 and April 19, 2020. The flow chart of our literature search is presented in **Figure 1**. Descriptive characteristics of the eligible studies are presented in **Supplementary Table 1**. Sixteen articles were excluded, because all the reported associations were also examined in a larger study with overlapping population. All the eligible studies were hospital-based observational studies. Six of the 88 eligible articles (7%) were conducted outside China (France^19^, Hong Kong^20^, Italy^21^, Singapore^22^, United States of America^23,24^).

**Figure 1.**
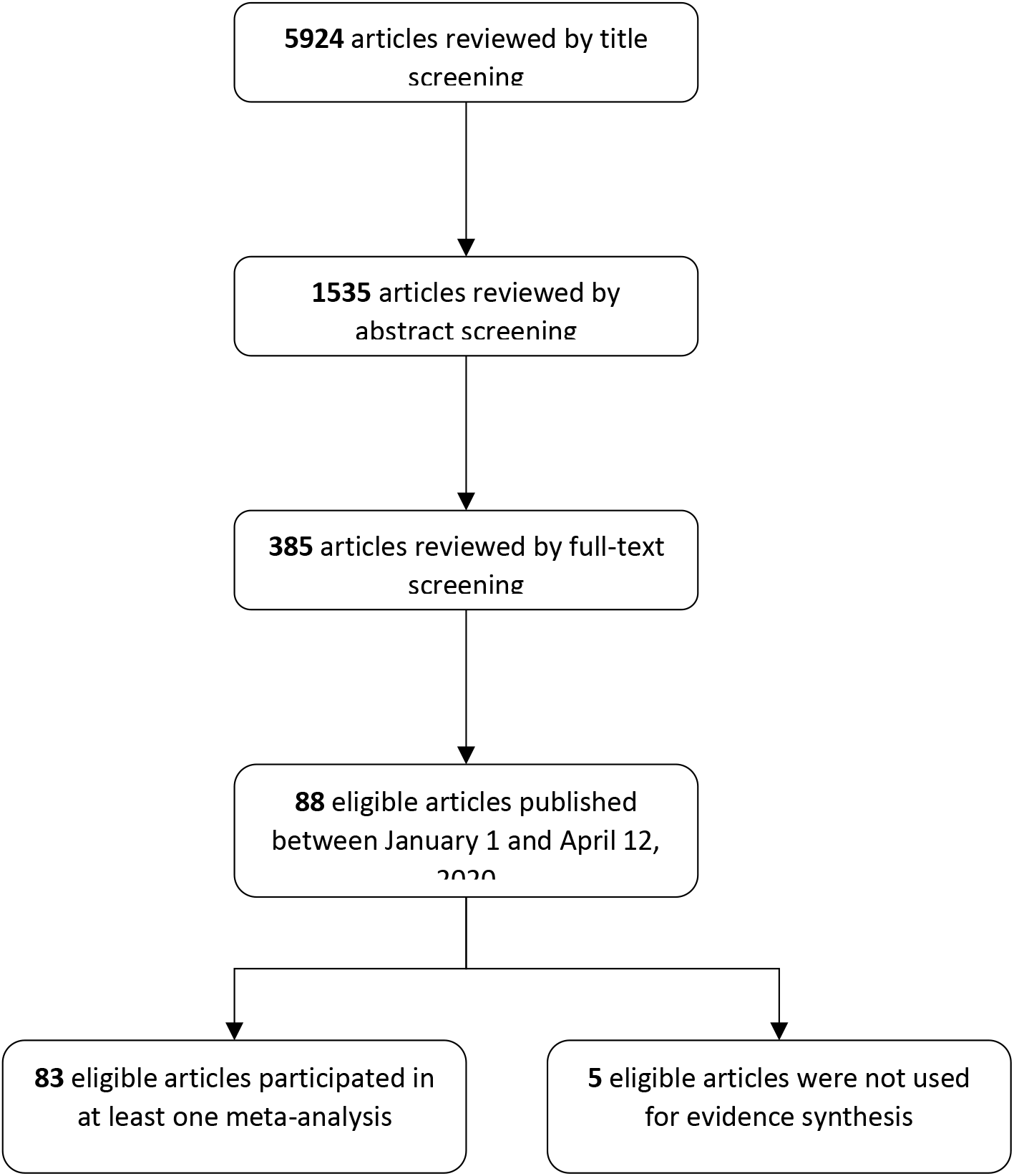
Flow chart of literature search

**Table 1.**
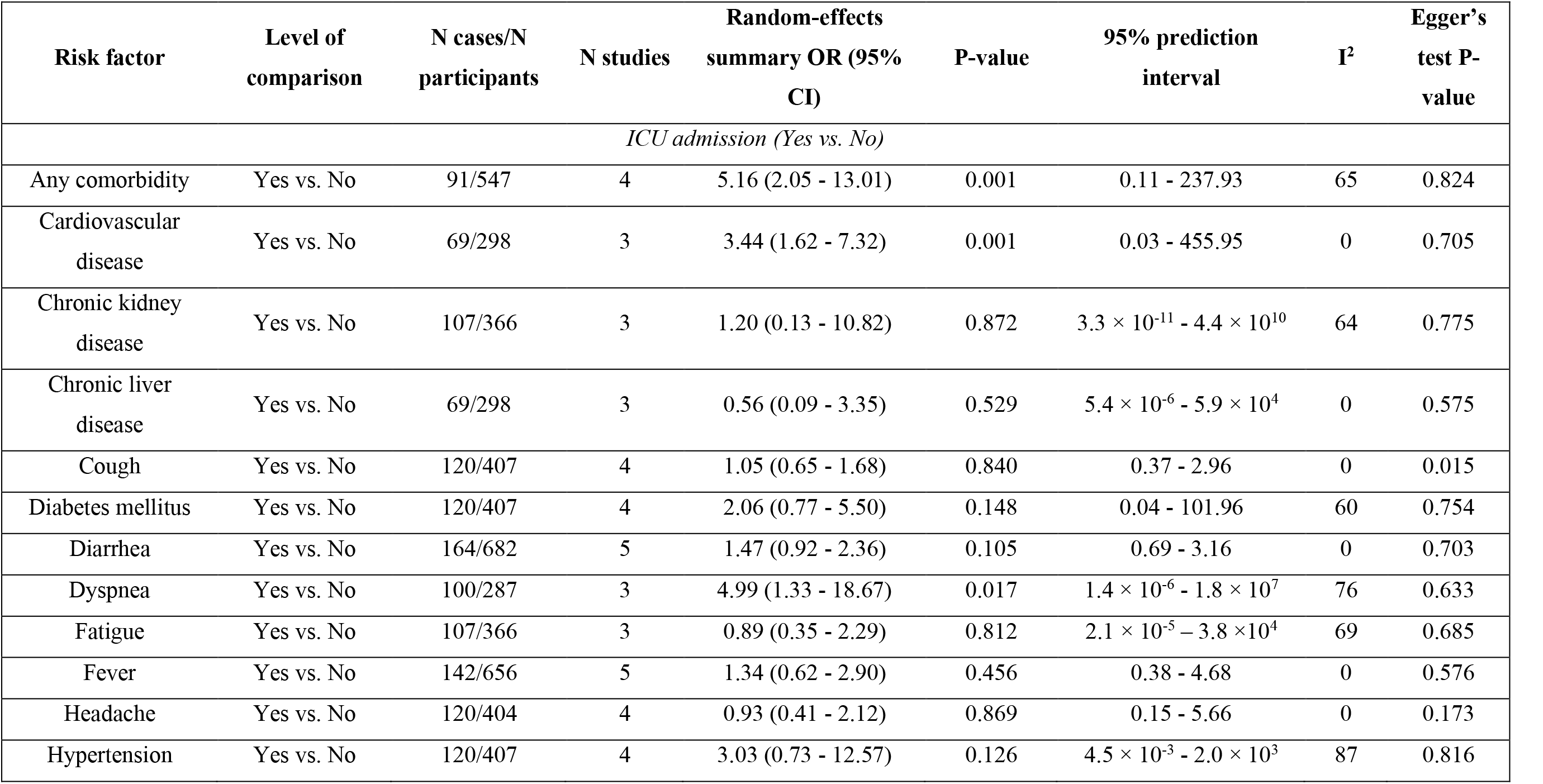
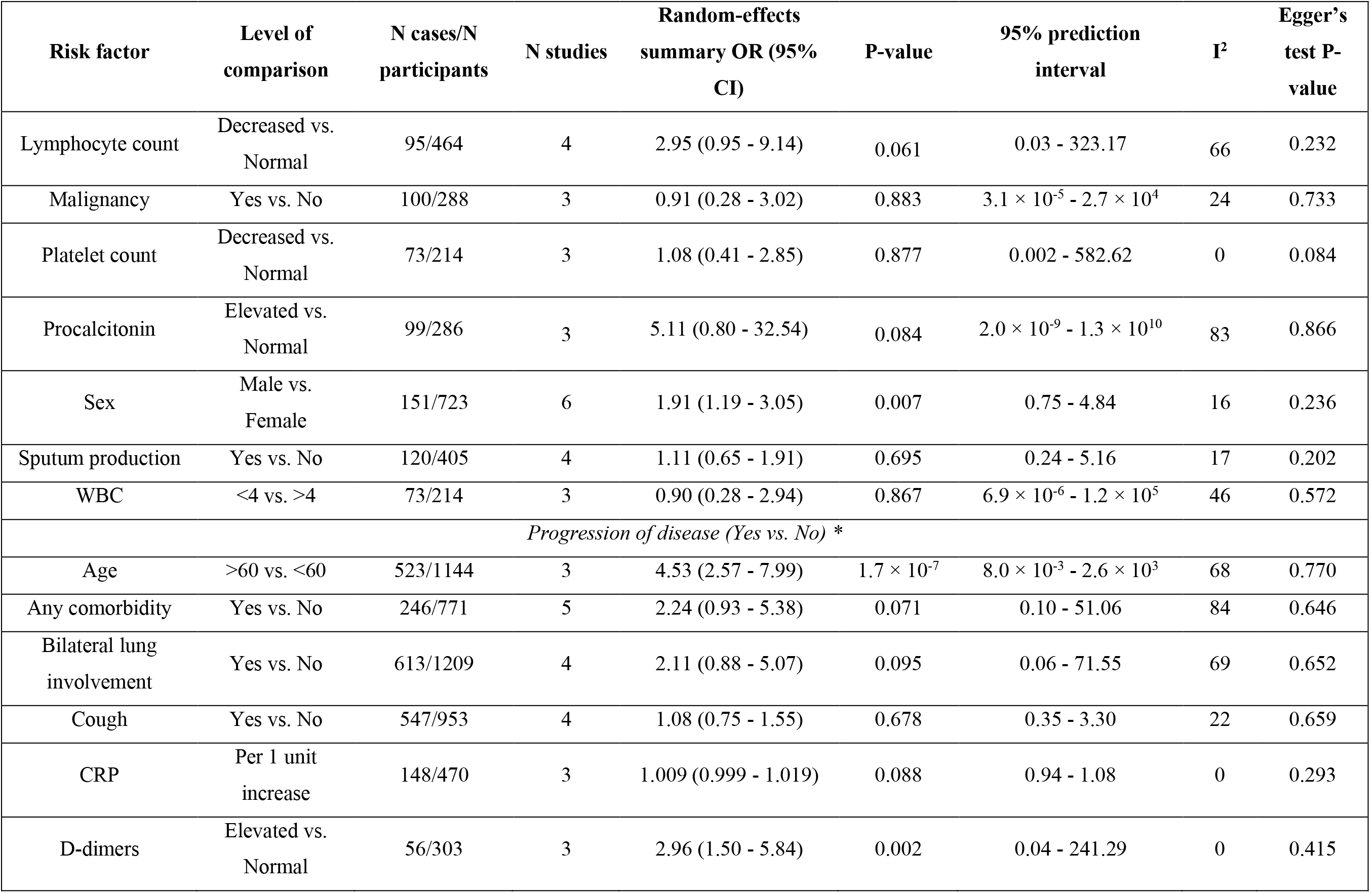
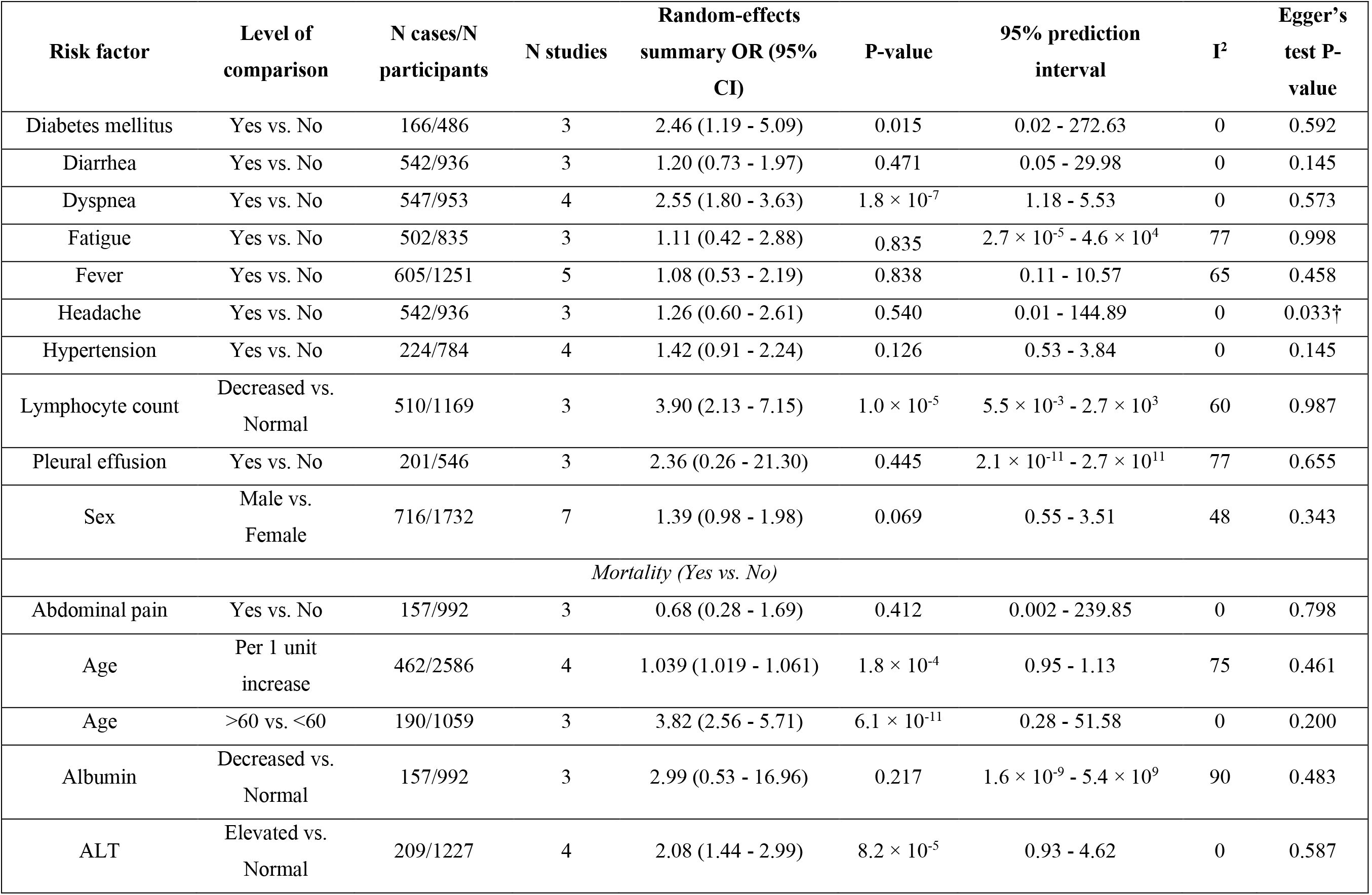
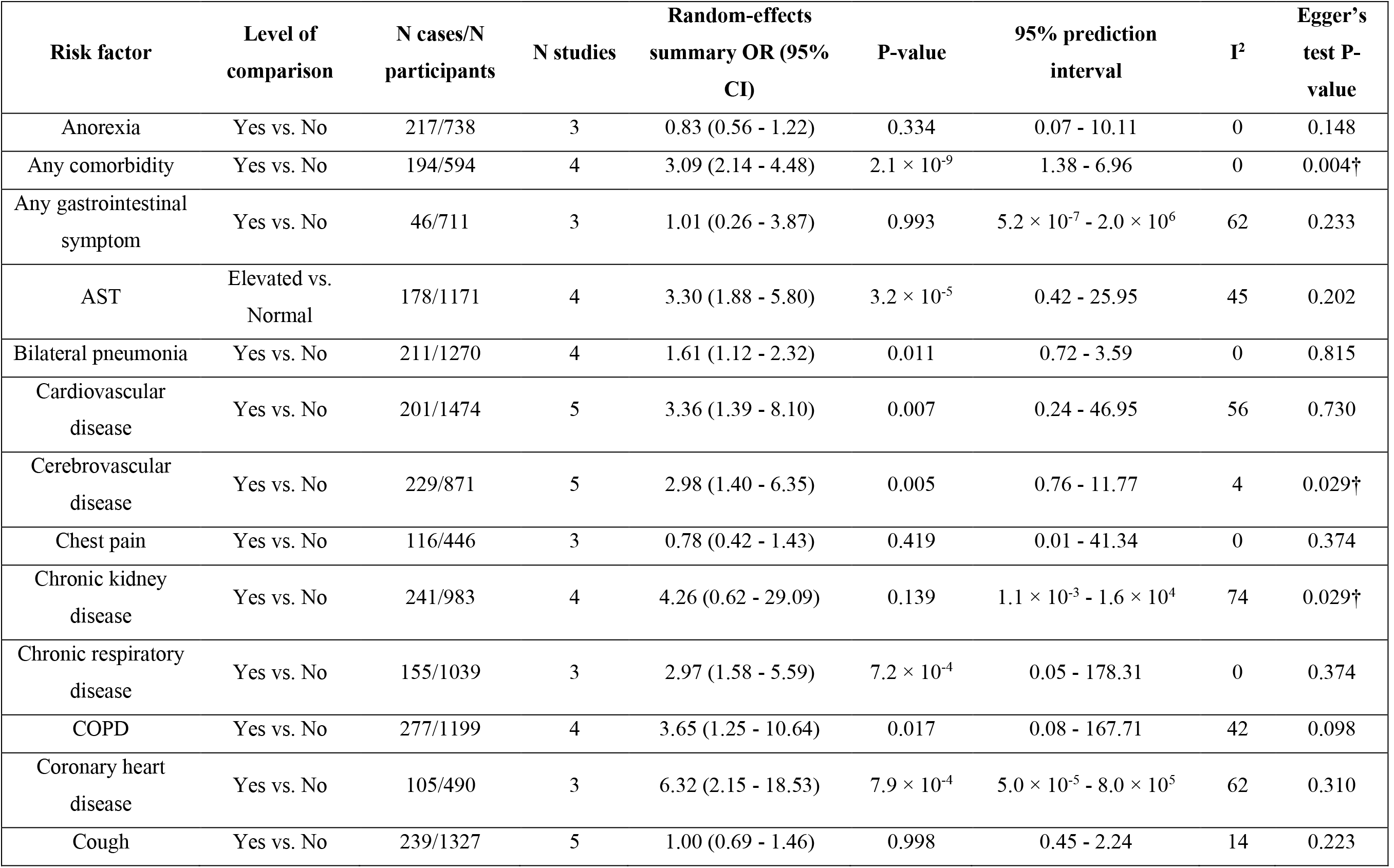
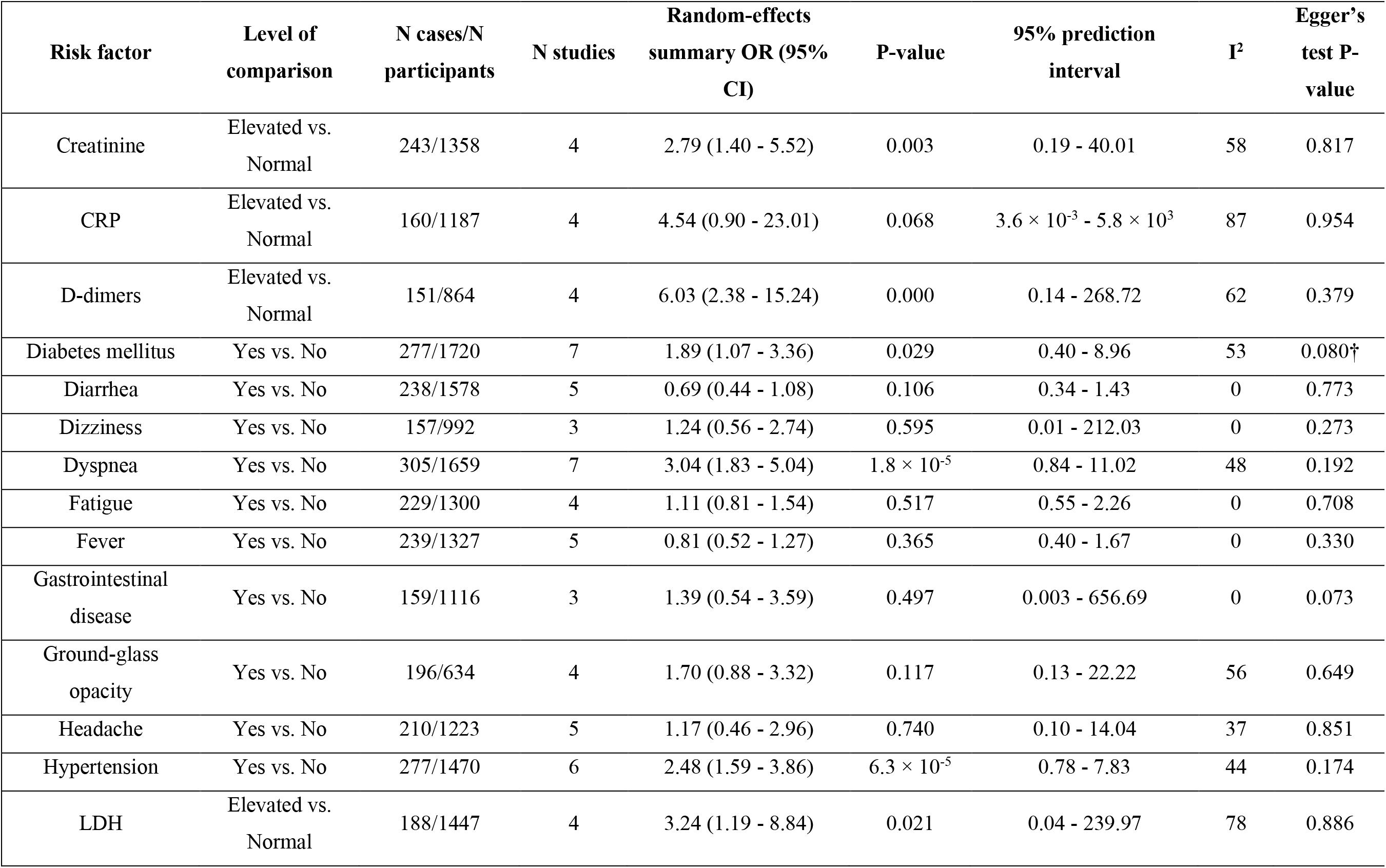
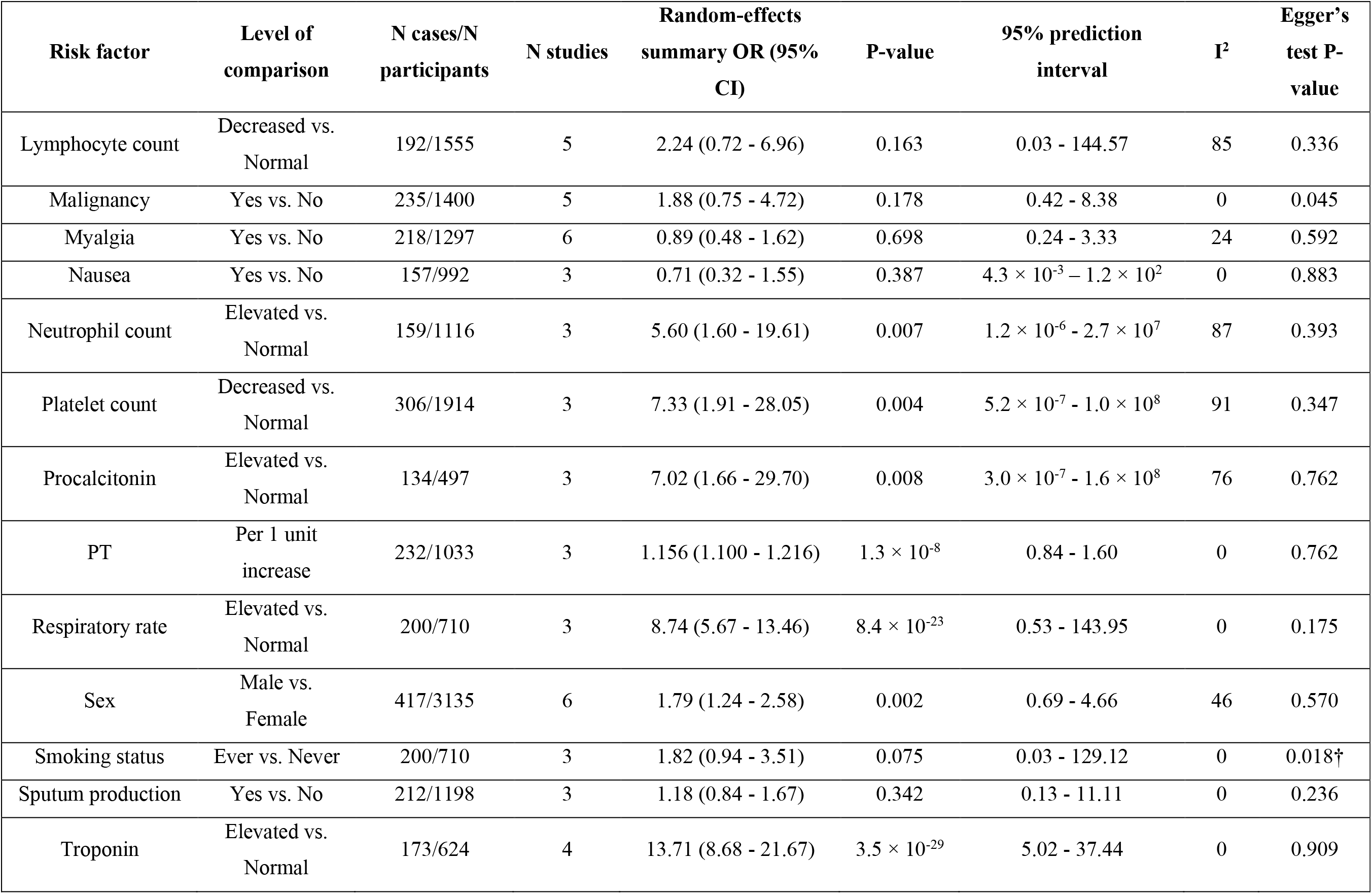
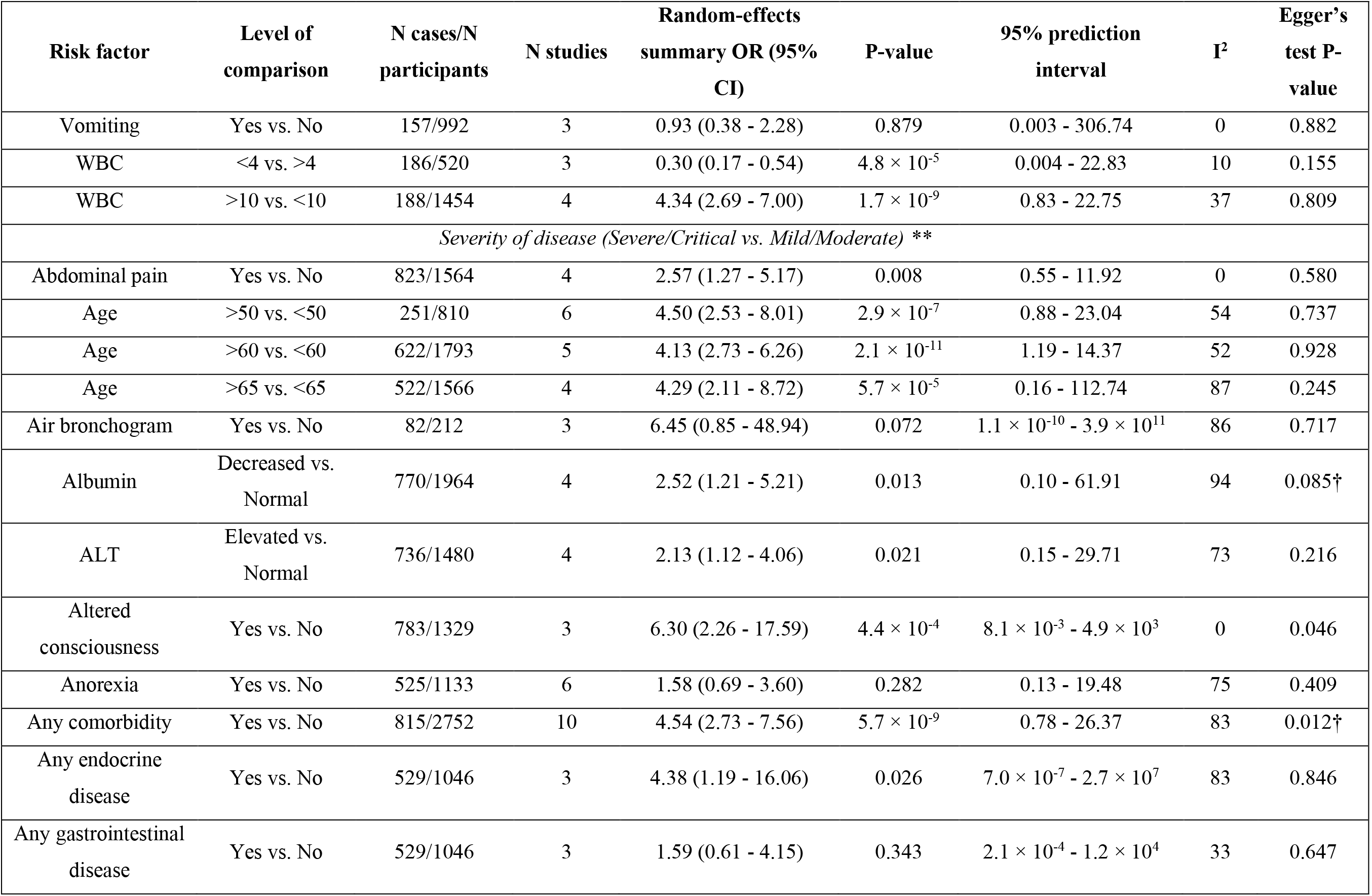
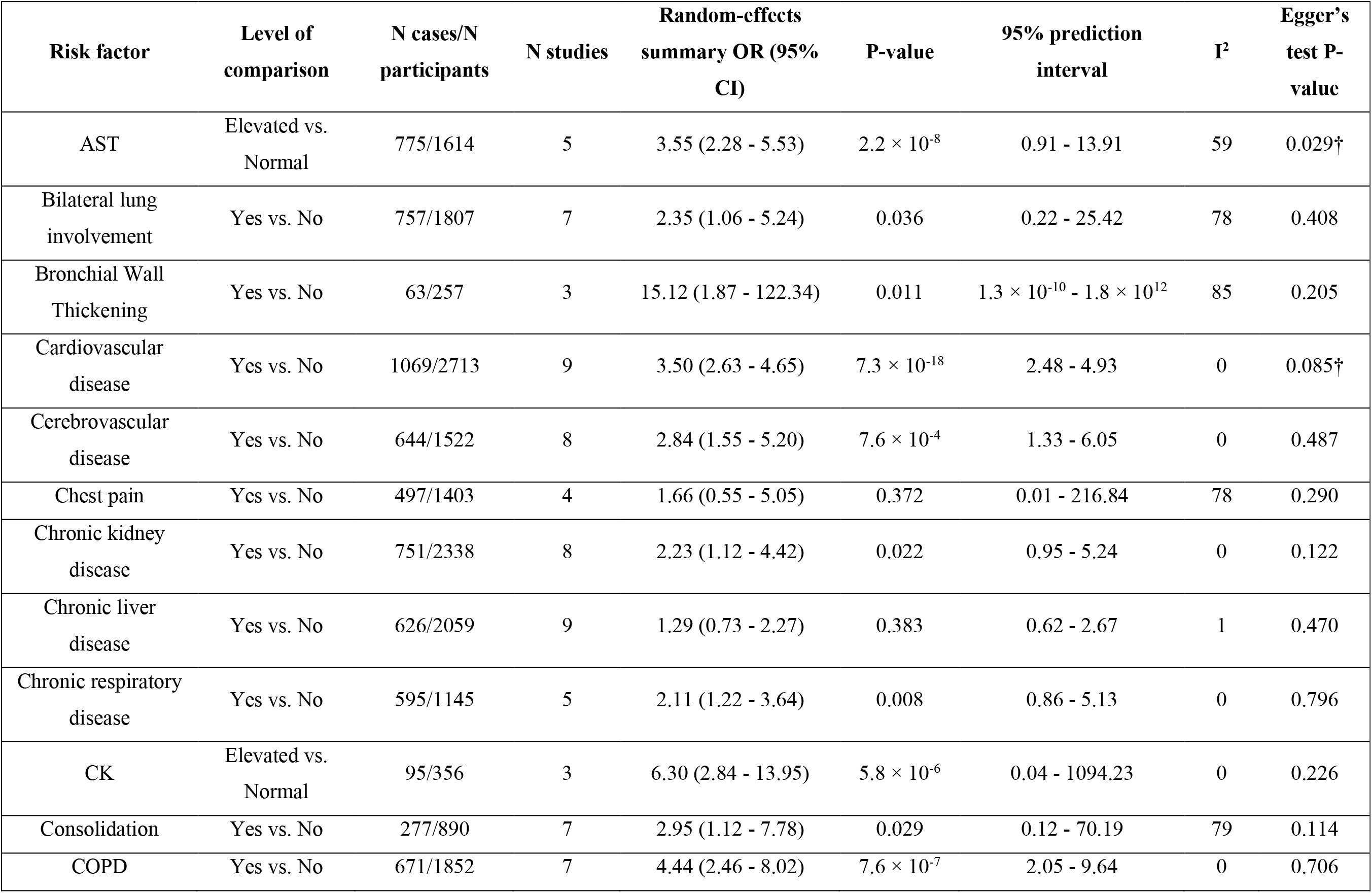
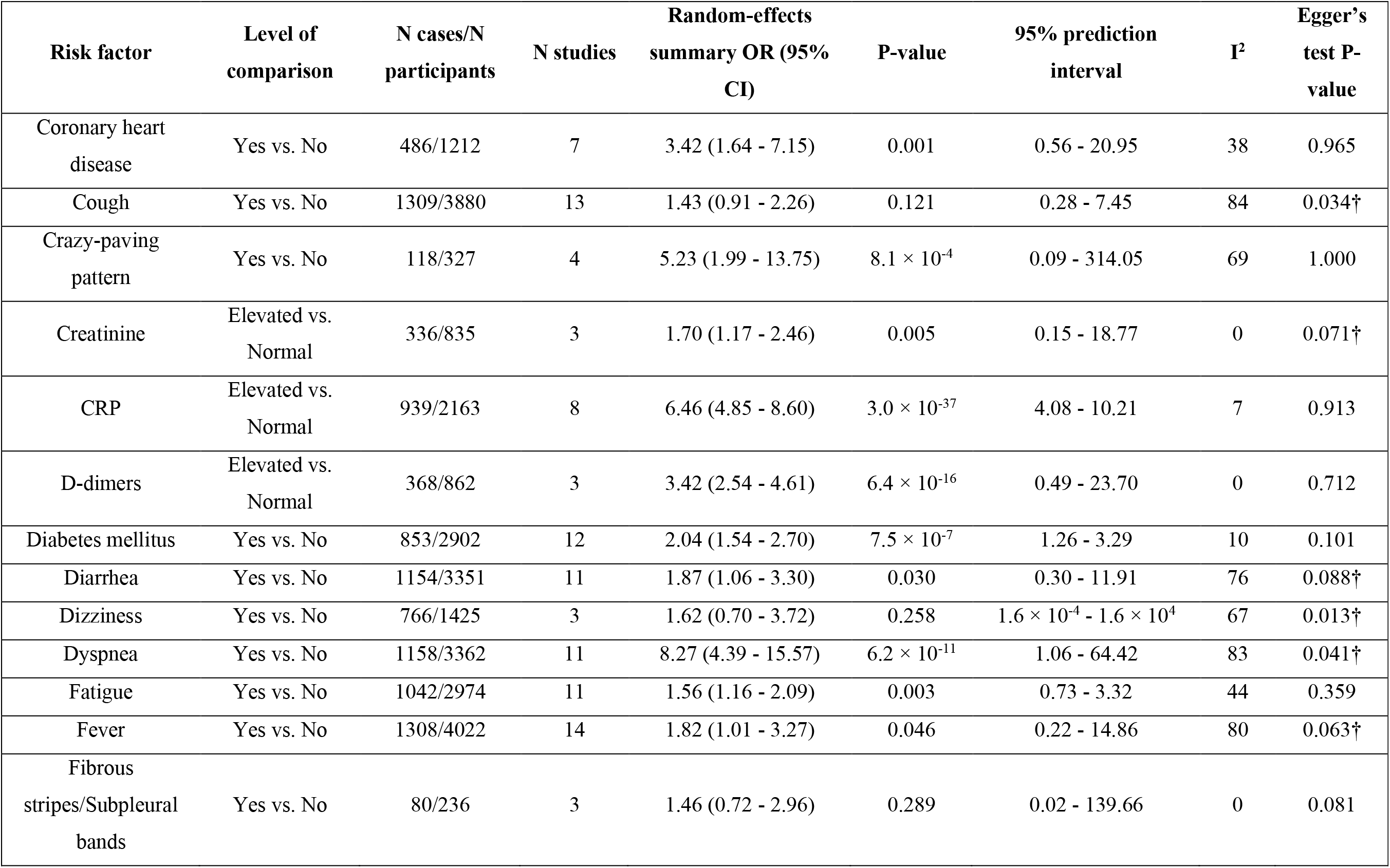
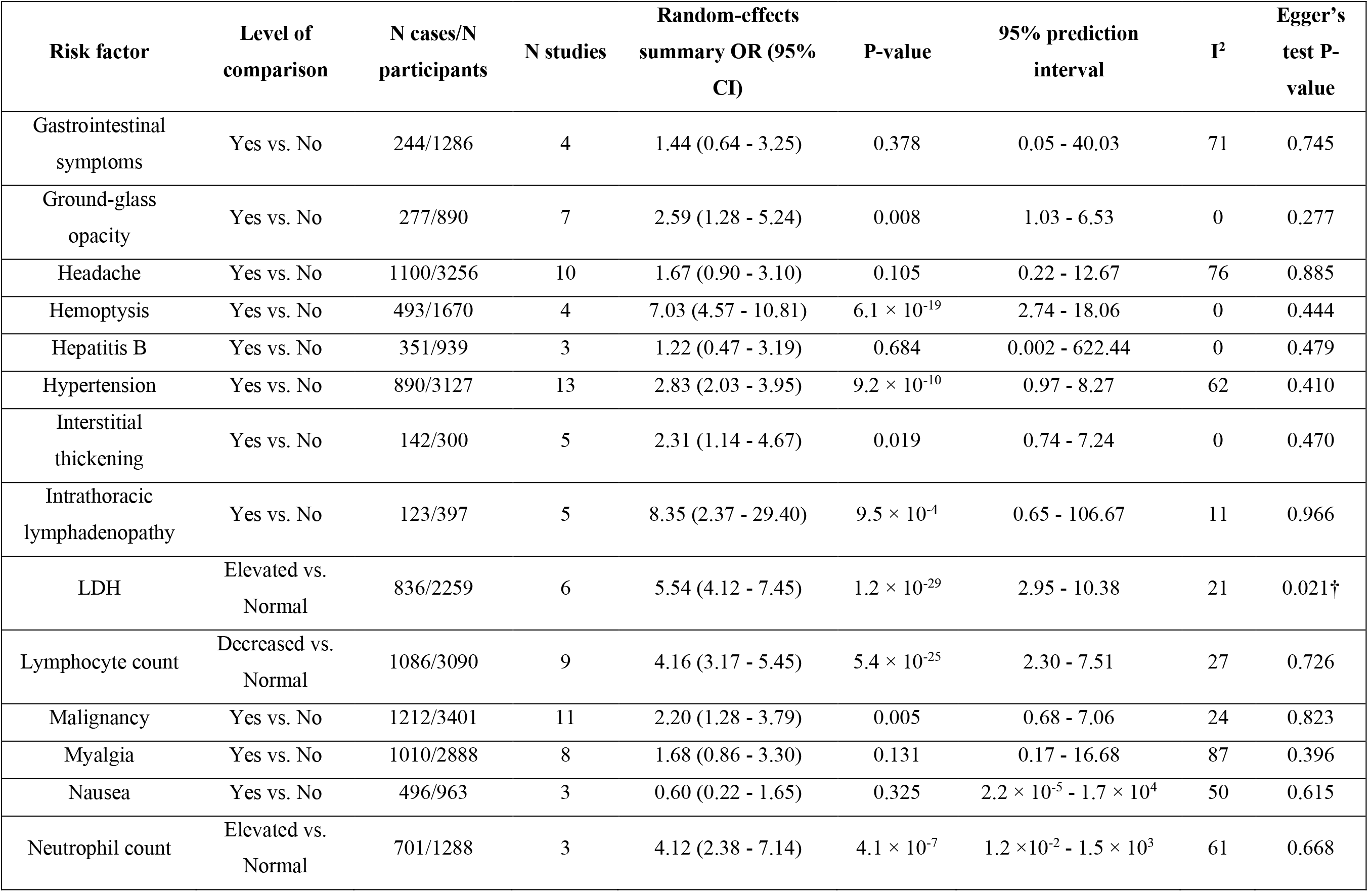
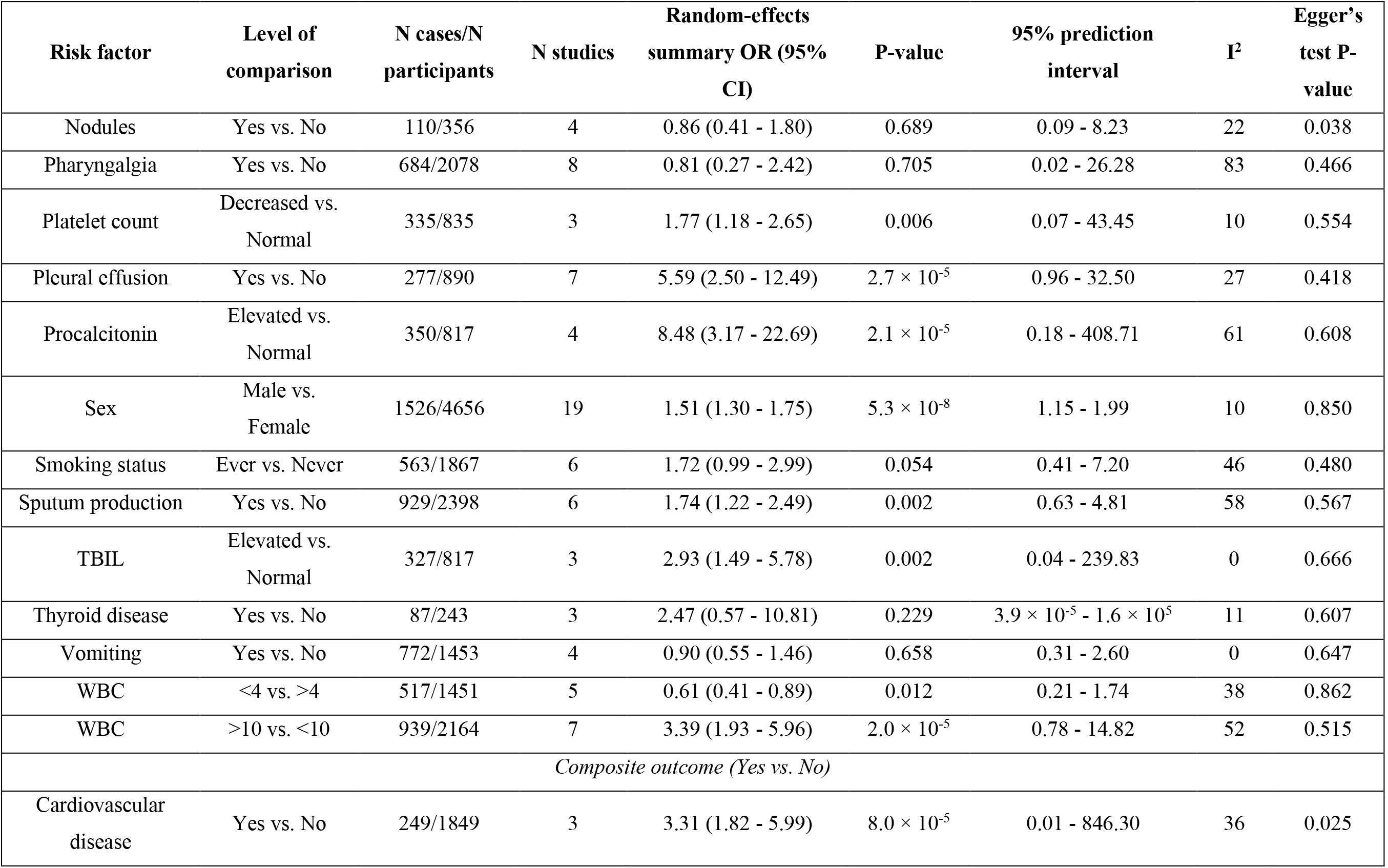
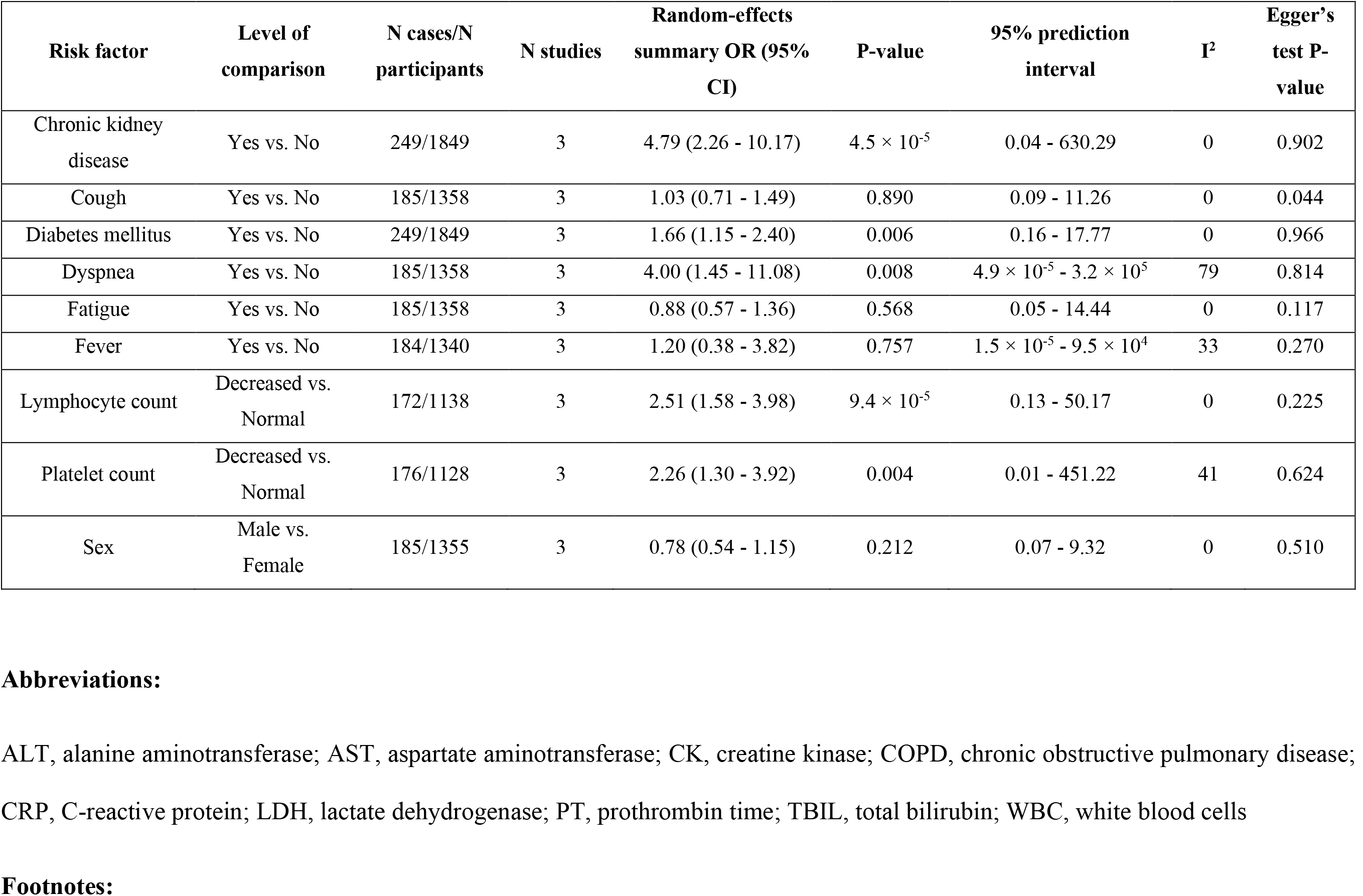
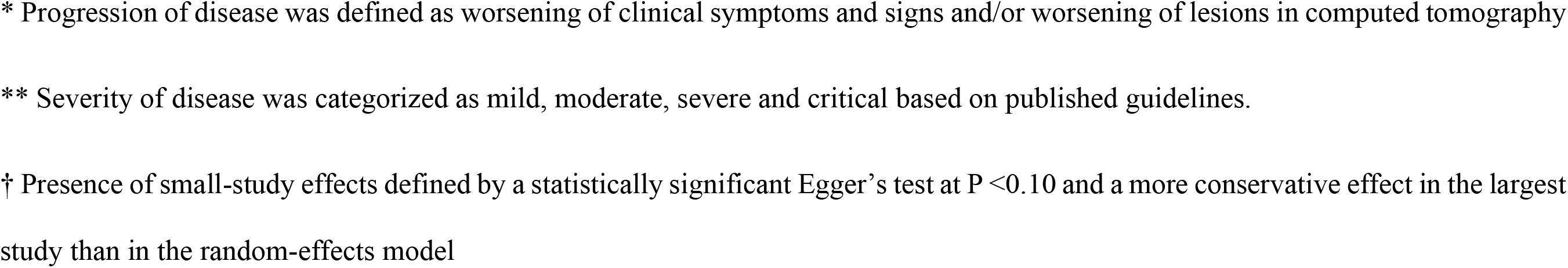
Meta-analyses of observational studies for risk factors of adverse clinical outcomes in patients with COVID-19 that were examined in at least 3 independent studies.

The outcomes that were examined in multiple articles were risk for severe COVID-19 (n = 86 meta-analyses), risk for mortality (n = 74 meta-analyses), risk for admission to ICU (n = 35 meta-analyses), risk for disease progression (n = 33 meta-analyses), risk for a composite outcome (n = 25 meta-analyses), and risk for invasive mechanical ventilation (n = 3 meta-analyses). A total of 256 meta-analyses were performed. For 4 outcomes (i.e., hospital admission^25,26^, acute respiratory distress syndrome^27^, severity of COVID-19 in pediatric population^28^, need for supplemental oxygen^29^), a meta-analysis was not performed, because each one of the reported associations for these outcomes was considered in only one study.

Overall, we identified 98 unique risk factors for adverse clinical outcomes in patients with COVID-19. We categorized these risk factors into five categories: biomarkers (n = 71 meta-analyses, 26 unique factors), comorbidities (n = 60 meta-analyses, 23 unique factors), imaging markers (n = 23 meta-analyses, 17 unique factors), demographic characteristics (n = 20 meta-analyses, 3 unique factors) and symptoms and clinical signs (n = 82 meta-analyses, 29 unique factors). The five meta-analyses with the largest number of studies considered sex (n = 19), fever (n = 14), cough (n = 13), hypertension (n = 13), and diabetes mellitus (n = 12) with severity of COVID-19.

The median number of studies per association was 3 (interquartile range, 2 – 4), whereas the median number of cases was 189 (interquartile range, 100 – 457) and the median number of participants was 818 (interquartile range, 442 – 1307). One hundred of 256 (39%) included more than 1000 patients with COVID-19. One hundred and twenty of 256 meta-analyses (47%) presented a statistically significant effect at P-value <0.05. Ninety-two of 256 meta-analyses (36%) presented large between-study heterogeneity (I^2^ > 50%). One hundred and fifty-seven of 256 meta-analyses (61%) included at least three independent studies, allowing for the estimation of 95% prediction interval and for the application of Egger’s test. Only fifteen of 157 meta-analyses (10%) presented a 95% prediction interval that excluded the null value. These associations examined the effect of any comorbidity and troponin on risk of mortality, the effect of dyspnea on risk of progression, and the effect of C-reactive protein (CRP), lactate dehydrogenase (LDH), lymphocyte count, cardiovascular disease, cerebrovascular disease, chronic obstructive pulmonary disease (COPD), diabetes mellitus, dyspnea, hemoptysis, age, sex and ground-glass opacity on risk of severe COVID-19. Also, 27 of 157 meta-analyses (17%) presented a statistically significant result in Egger’s test at P-value <0.10. Seventeen of these 27 meta-analyses presented a more conservative effect in the largest study than in random-effects meta-analysis, indicating the presence of small-study effects. These associations examined the effect of headache on risk of disease progression, the effect of any comorbidity, cerebrovascular disease, chronic kidney disease, diabetes mellitus, and smoking status on risk of mortality, and the effect of albumin, aspartate aminotransferase (AST), creatinine, LDH, any comorbidity, cardiovascular disease, cough, diarrhea, dizziness, dyspnea, and fever on risk of severe COVID-19. The results of the meta-analyses for the 256 associations are presented in **Table 1** and **Supplementary Table 2**.

Of the 120 statistically significant meta-analyses at P-value <0.05, seven meta-analyses included more than 1000 patients with COVID-19 in at least 3 independent studies, had a P-value <0.005 in random-effects model, low to moderate between-study heterogeneity, 95% prediction interval excluding the null value and absence of small-study effects. These meta-analyses examined the effect of serum CRP (OR,6.46; 95% CI, 4.85 – 8.60), lymphocyte count (OR, 4.16; 95% CI, 3.17 – 5.45), cerebrovascular disease (OR,2.84; 95% CI, 1.55 – 5.20), COPD (OR, 4.44; 95% CI, 2.46 – 8.02), diabetes mellitus (OR,2.04; 95% CI, 1.54 – 2.70), hemoptysis (OR, 7.03; 95% CI, 4.57 – 10.81), and sex (OR,1.51; 95% CI, 1.30 – 1.75) on risk of severe COVID-19. Seventeen additional meta-analyses fulfilled the aforementioned characteristics with the exception of a 95% prediction interval that included the null value. These meta-analyses assessed the effect of alanine aminotransferase (ALT), AST, prothrombin time, chronic respiratory disease, hypertension, dyspnea, age, sex, elevated white blood cells on risk of mortality, the effect of coronary heart disease, malignancy, altered consciousness, and fatigue on risk of severe COVID-19, and the effect of chronic kidney disease, lymphocyte count, platelet count, and cardiovascular disease on risk of a composite outcome. A forest plot of these 24 associations is presented in **Figure 2**.

**Figure 2.**
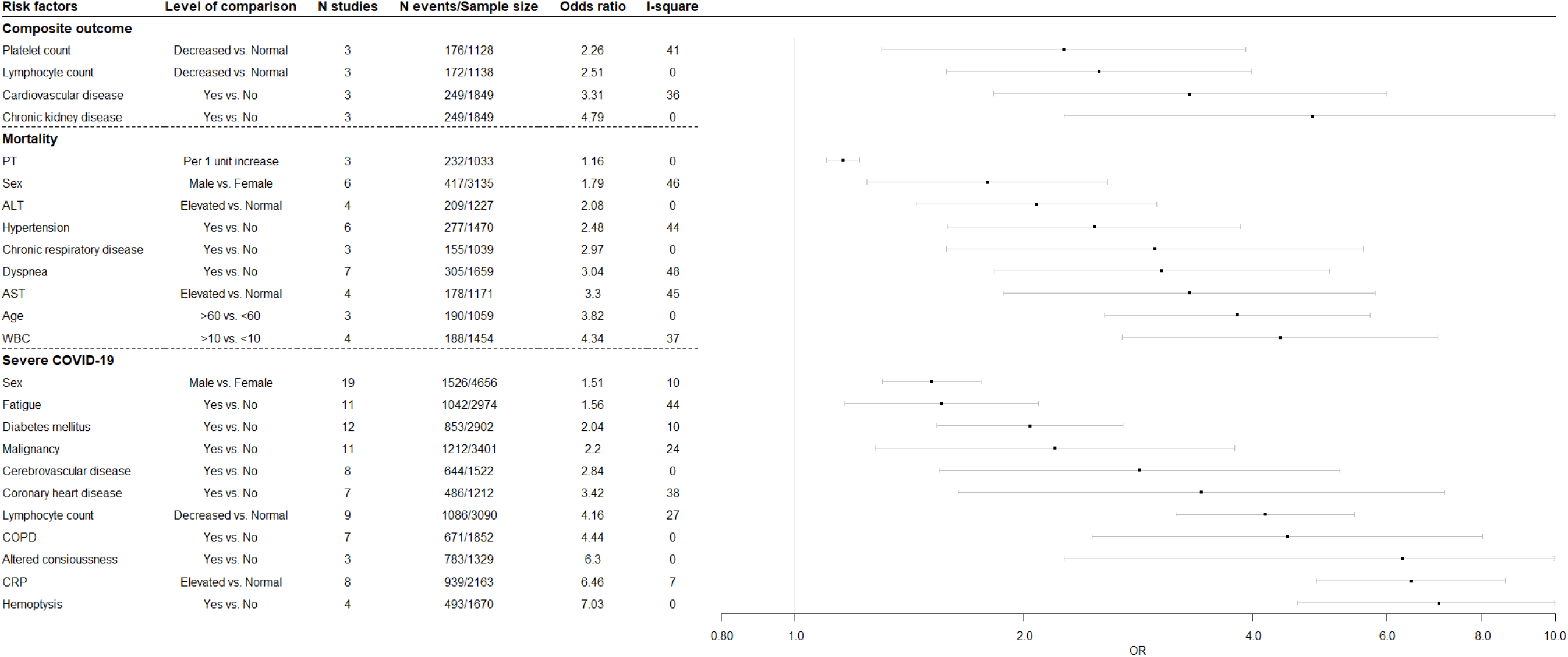
Forest plot of the 24 statistically significant (P-value <0.005) associations that had more than 1000 COVID-19 patients, I^2^ <50% and at least 3 independent studies.

## 4 DISCUSSION

We conducted a systematic review and meta-analysis to present an overview of all the risk factors associated with adverse clinical outcomes in COVID-19 patients. In the present effort, we considered more than 250 associations that were related to the effect of various risk factors on adverse clinical outcomes for COVID-19. Almost half of these associations presented a nominally significant effect, and only 10 associations presented the strongest evidence in terms of sample size, statistical significance, consistency and lack of small-study effects.

Our literature search identified a multitude of articles examining the effect of various clinical characteristics on clinical course of COVID-19. However, detailed examination of these articles revealed a great number of studies examining the same associations in overlapping populations, and, therefore, we scrutinized eligible articles to ensure we include only independent populations in each meta-analysis. For example, a large nationwide study from China containing 1099 participants reported various characteristics for an association with a composite clinical adverse outcome and mortality.^30^ Due to overlap, this study was only considered in meta-analysis consisted of non-Chinese studies exclusively. This was because when other studies from various provinces of China examined an association the summary sample was greater than the sample of the nation-wide study.

There is a wide spectrum of clinical presentations for COVID-19 patients.^31^ The clinical features that were associated with adverse clinical events were mainly respiratory symptoms or non-specific flu-like symptoms. Dyspnea showed a significant association with all the adverse outcomes that we examined. It can be deduced that patients experiencing shortness of breath might have pneumonia and considerable lung damage, and dyspnea might be a symptom of worsening respiratory failure. This rational could also explain the observed association between respiratory rate and risk of mortality and altered consciousness and severity of disease, as both symptoms might result from deteriorating respiratory failure. Hemoptysis and sputum production were also linked with an increased risk for severe COVID-19 and may be proxies of extensive lung damage from SARS-CoV-2 or bacterial co-infection or symptoms of preexisting chronic lung disease such as bronchiectasis, and COPD. Also, hemoptysis could be a result of endothelial disruption and pulmonary vessel coagulopathy, which have been described in literature.^32^ This finding is further corroborated by the association we unveiled between d-dimers and various adverse outcomes, as well as observational studies depicting increased thrombotic incidents in COVID-19 patients.^33^ Symptoms typical in flu-like illness that originate outside the respiratory system and were linked with severe COVID-19 were fever, fatigue, diarrhea, and abdominal pain. Symptoms that have a negative impact on disease course could potentially be useful during patient assessment, i.e. they might be considered “red flags” that pinpoint patients being at high risk for a negative prognostic outcome and requiring enhanced monitoring or treatment.^34^

Our results depicted that age is a risk factor related to severe COVID-19, progression of disease and mortality. Also, a dose-response association was observed between age and risk of mortality. However, it cannot be assumed that this finding is causal given that several chronic diseases may be confounders of this association. This finding is in accordance with our analysis indicating that major comorbid disorders of late adult life, such as cardiovascular and cerebrovascular disease, diabetes mellitus and COPD, are also markers associated with various adverse clinical events. Also, sex was identified as a statistically significant risk factor, presenting a 1.5-fold increase in the risk of severe COVID-19 and, a 1.8-fold increase in the risk of mortality and a 1.9-fold increase in the risk of ICU admission in male compared to female patients.

Presence of chronic comorbid disorders also increased the risk of severe COVID-19, ICU admission and mortality. There is published evidence that SARS-CoV-2 causes systemic inflammation inducing a cytokine storm and may inflict multi-organ damage outside the respiratory system as a result such as myocardial injury, acute kidney damage, liver injury.^33^ Patients with pre-existing chronic disorders of organs damaged by SARS-CoV-2 may be more prone to organ insufficiency for that matter. This might be the case for cardiovascular disease, coronary heart disease, hypertension, chronic kidney disease and cerebrovascular conditions. These conditions were associated with elevated risk of various adverse clinical outcomes in COVID-19 patients. The association of hypertension with poor prognosis might be a result of the various systemic complications of hypertension. Another potential mechanism implicates the angiotensin converting enzyme, which is overexpressed in patients with hypertension and cardiovascular diseases.^35^ This enzyme is a functional receptor of SARS-CoV-2 and it is hypothesized that it could be responsible for acute myocardial injury inflicted by SARS-CoV-2.^35^ Also, patients with COPD and other chronic lung diseases had an increased risk for severe COVID-19 and mortality. These patients have non-reversible structural lung abnormalities and lung function impairment.^17^ They may have bacterial colonization, impaired immune defense and mucocilliary clearance of the lower respiratory tract, which might impair viral clearance of SARS-CoV-2. At the same time, COPD patients exhibit chronic respiratory failure and diminished respiratory reserve and infection by any virus may instigate an exacerbation of the disease. Furthermore, patients with diabetes mellitus, and malignancy presented worse prognosis. Both these patients have a compromised immune system, which disrupts patients’ response to a viral agent.^36,37^ There is published evidence that all the aforementioned comorbid diseases are also important factors that impact influenza illness severity.^38^

The identification of a serum biomarker to help individual risk stratification for each patient with COVID-19 could offer additional clinical insights.^31^ In our analysis, a total of 16 serum biomarkers presented a significant effect on adverse clinical outcomes based on meta-analyses of at least three independent studies. Decreased lymphocyte count was associated with higher risk of progression of disease, and presentation with severe COVID-19 and is a laboratory feature used for disease diagnosis.^39^ Elevated leukocyte count, CRP and procalcitonin are inflammatory markers, and they had a detrimental effect on disease course.^31^ Moreover, serum levels of creatine kinase (CK) was associated with severity of disease, whereas LDH was associated with both severity of disease and mortality. Both are biomarkers increased from tissue damage, for LDH originating in any organ, while for CK originating mainly in the heart, skeletal muscle and kidney. For the same reason troponin might be a surrogate market of myocardial injury caused by SARS-CoV-2 and therefore is associated with mortality. Also, dysregulation of liver and renal function, defined by abnormalities in serum ALT, AST, total bilirubin and albumin in the first case and increased serum creatinine in the second case, were associated with an elevated risk of adverse clinical events. Furthermore, the presence of blood coagulation disorders in patients with COVID-19 has a significant clinical impact, as confirmed by the meta-analyses assessing platelet count, d-dimers and prothrombin time with adverse clinical outcomes.^32^

Moreover, several findings from computed tomography of chest were linked with unfavorable clinical endpoints in patients with COVID-19. Of note, imaging features of SARS-CoV-2 significantly overlap with radiological findings observed in infection by other coronaviruses causing the Middle East Syndrome and Severe Acute Respiratory Syndrome.^40^ Bilateral lung involvement increased the risk of both severe COVID-19 and mortality. Radiological patterns that were associated with severity of COVID-19 included ground glass opacities, consolidation, crazy paving and interstitial thickening. These findings might be a result of a more extensive inflammation of the lungs. Additional radiological findings that were associated with severe COVID-19 were, bronchial wall thickening, and intrathoracic lymphadenopathy. These may be a result of prolonged inflammation which leads to structural long-term sequelae such as pulmonary interstitial fibrosis and bronchiectasis, as described after H1N1 influenza pneumonia and Severe Acute Respiratory Syndrome.^41,42^ Presence of pleural effusion was associated with severe COVID-19. There is published evidence that pleural effusion was a poor prognostic factor in MERS-CoV and H5N1 avian influenza patients.^43^ Pleural effusion could be an attribute of pleural inflammation from the SARS-CoV-2 or bacterial co-infection or exacerbation of pre-existing heart failure.

A systematic review of prediction models focusing on prognosis of COVID-19 patients retrieved 10 published models.^44^ The majority of these models included age, sex, LDH, CRP, lymphocytes, radiological markers of disease burden and presence of comorbidities among their predictors. Based on this finding, variables included in these models overlap greatly with the risk factors that resulted from our study. Also, the additional risk factors that were identified by our study could be considered as potential predictors in future updates of the available prognostic models.

Some limitations should be considered for the interpretation of our findings. A majority of the effect estimates are unadjusted odds ratios, indicating that potential confounding cannot be excluded. Moreover, a majority of the studies included in our meta-analyses were based on samples from China. There is need for independent observational studies to examine the external validity of our findings. Also, the observed association of some biomarkers with severity and progression of disease might be a result of reverse causation. This means that the observed differences in the biomarkers could be attributed to the natural history of COVID-19. Furthermore, a third of the meta-analyses presented large between-study heterogeneity, but sources of heterogeneity could not be explored due to the small number of observational studies in each meta-analysis. Potential sources of heterogeneity include different methods of risk factor measurement, varying inclusion criteria between studies as well as random error due to the small sample size of the studies.

Our article systematically identified all the published observational studies examining risk factors for adverse clinical outcomes in patients with COVID-19. The systematic review was further complemented by meta-analysis on more than 250 associations. The risk factors that presented the strongest epidemiological evidence were related to age, sex, respiratory and flu-like symptoms and clinical signs, major comorbidities, pleural effusion, inflammatory markers, biomarkers of coagulation and tissue damage. However, these findings were mainly based on unadjusted effect estimates, indicating that they might not be genuine, and confounding could influence some of the them. For this reason, our findings should not be interpreted as causal associations. Future studies should present adjusted effect estimates for age, sex and other potential confounders. Also, as more observational studies accumulate on risk factors, it is likely that certain factors studied in a larger sample might surface as statistically significant, and sources of heterogeneity would become clearer.

## Data Availability

Data are available upon reasonable request.

